# Saliva as a Candidate for COVID-19 Diagnostic Testing: A Meta-Analysis

**DOI:** 10.1101/2020.05.26.20112565

**Authors:** László Márk Czumbel, Szabolcs Kiss, Nelli Farkas, Iván Mandel, Anita Hegyi, Ákos Nagy, Zsolt Lohinai, Zsolt Szakács, Péter Hegyi, Martin C. Steward, Gábor Varga

## Abstract

**Objectives:** Our aim was to conduct a meta-analysis on the reliability and consistency of SARS-CoV-2 viral RNA detection in saliva specimens.

**Methods:** We reported our meta-analysis according to the Cochrane Handbook. We searched the Cochrane Library, Embase, Pubmed, Scopus, Web of Science and clinical trial registries for eligible studies published between 1 January and 25 April 2020. The number of positive tests and total number of conducted tests were collected as raw data. The proportion of positive tests in the pooled data were calculated by score confidence interval estimation with the Freeman-Tukey transformation. Heterogeneity was assessed using the I^2^ measure and the ***χ**^2^* test.

**Results:** The systematic search revealed 96 records after removal of duplicates. 26 records were included for qualitative analysis and 5 records for quantitative synthesis. We found 91% (95%CI = 80%-99%) sensitivity for saliva tests and 98% (95%CI 89%-100%) sensitivity for nasopharyngeal swab (NPS) tests in previously confirmed COVID-19 infected patients, with moderate heterogeneity among studies. Additionally, we identified 18 registered, ongoing clinical trials on saliva-based tests for detection of the virus.

**Conclusion:** Saliva tests offer a promising alternative to NPS for COVID-19 diagnosis. However, further diagnostic accuracy studies are needed to improve their specificity and sensitivity.

## Introduction

COVID-19 caused by SARS-CoV-2 is a serious and potentially deadly disease. Globally, as of 5 May 2020, there have been 3,489,053 confirmed cases of COVID-19, including 241,559 deaths, reported to WHO on 5 May 2020 (World Health, 2020b). Early diagnosis and isolation of infected individuals will play an important role in stopping the further escalation of the pandemic.

At present, nasopharyngeal swabbing, followed by reverse transcription of the extracted RNA and quantitative PCR (RT-qPCR) is the gold standard for detection of SARS-CoV-2 infection (Lippi, Simundic, & Plebani, 2020). Specimen collection currently requires trained personnel (World Health, 2020a), thus exposing medical staff to a higher risk of infection (Kim, Yun, Kim, Park, Cho, Yoon, Nam, Lee, Cho, & Lim, 2017). It is not always successful at the first attempt, and shortages of swabs and protective equipment are frequently reported (Lippi et al., 2020). Additionally, mass testing requires an increased number of trained personnel at specimen acquiring sites. Consequently, the nasopharyngeal swab (NPS) collection method is causing an economic and logistic burden on healthcare systems. Additionally, nasopharyngeal swabbing causes serious discomfort to the patients (Li, Liu, Yu, Tang, & Tang, 2020) and there are several contraindications, such as coagulopathy or anticoagulant therapy and significant nasal septum deviation (Sri Santosh, Parmar, Anand, Srikanth, & Saritha, 2020). Clearly, there is a need for a simple, less invasive method that also reduces the risk to healthcare personnel.

One candidate for non-invasive specimen collection is saliva. The saliva secreted by salivary glands contains water, electrolytes, mucus, and digestive and protective proteins (Dawes & Wong, 2019; Humphrey & Williamson, 2001; Varga, 2015). But whole saliva is a mixture of glandular secretions, gingival crevicular fluid, serum, expectorated airway surface liquid and mucus, epithelial and immune cells from the oral mucosa and upper airways, and oral microbes and viruses (Miller, Foley, Bailey, Campell, Humphries, Christodoulides, Floriano, Simmons, Bhagwandin, Jacobson, Redding, Ebersole, & McDevitt, 2010). Despite its heterogeneous origins, this mixed fluid is widely used as a diagnostic tool to identify various oral and systemic conditions ((Dawes & Wong, 2019; Keremi, Beck, Fabian, Fabian, Szabo, Nagy, & Varga, 2017). These already include viral infections such as dengue, West Nile, chikungunya, Ebola, Zika and Yellow Fever, and also the recently emerged coronaviruses responsible for severe acute respiratory syndrome (SARS) and Middle East respiratory syndrome (MERS) (Niedrig, Patel, El Wahed, Schadler, & Yactayo, 2018).

Since early January 2020, several papers have been published on the possible use of saliva as a specimen for detecting SARS-CoV-2 in the diagnosis of COVID-19. Until now there has been no systematic review or meta-analysis of this topic. Therefore, our aim was to conduct a meta-analysis to overcome the limitations of small sample sizes in the individual studies in order to estimate the diagnostic sensitivity of saliva-based detection of the disease. We also aimed to summarize the study protocols which have been registered in clinical trial registries to investigate saliva-based COVID-19 identification in the future.

## Materials and methods

### Protocol and registration

The reporting of our meta-analysis follows the guidelines of the Preferred Reporting Items for Systematic Reviews and Meta-Analyses (PRISMA) (Moher, Shamseer, Clarke, Ghersi, Liberati, Petticrew, Shekelle, Stewart, & Group, 2015). The PRISMA checklist for our work is available in the supporting information (Table S1). We registered our meta-analysis protocol in the OSF (Open Science Framework by Center for Open Science) registries on 23 April 2020 (https://osf.io/3ajy7).

#### Deviation from the registered protocol

Studies eligible according to our inclusion criteria did not present sufficient raw data to complete 2×2 contingency tables. True positive, true negative, false positive and false negative values were not generally available, thus sensitivity and specificity could not be separately calculated. Instead, positive event rates were pooled for statistical analysis. Details of the analysis are described in section *Summary measures and synthesis of results*.

### Eligibility criteria

We included records if they have met the following eligibility criteria: 1) records published in scientific journals or clinical trial registry; 2) patients diagnosed with COVID-19; 3) index test: saliva specimens with PCR diagnostics for detecting SARS-CoV-2; 4) reference standard (comparator test): NPS specimens with PCR diagnostics for detecting SARS-CoV-2; 5) records written in English or available in English translation. Exclusion criteria: 1) publications with no primary results such as reviews, guidelines and recommendations; 2) publications dated before 1 Januaryand after 25 April, 2020; 3) gray and black literature.

### Search strategy

A systematic search in English language filtering for records published after 1 January 2020 was performed in five different major electronic databases (Cochrane Library, Embase, PubMed, Scopus, Web of Science) and also in five clinical trial registers (ClinicalTrial.gov, EU Clinical Trials Register, NIPH Clinical Trial Search, ISRCTN Registry, ANZCTR Registry). The last update of our systematic search was performed on 25 April 2020. Cited and citing papers of the relevant studies were screened for further eligible studies.

The following key words were applied to each database to identify eligible records: *(COVID 19 OR COVID19 OR Wuhan virus OR Wuhan coronavirus OR coronavirus OR 2019 nCoV OR 2019nCoV OR 2019-nCoV OR SARS CoV-2 OR SARS-CoV-2 OR NCP OR novel coronavirus pneumonia OR 2019 novel coronavirus OR new coronavirus) AND (saliva)*.

### Study selection

We used EndNote X9.3.3 reference manger to organize records. After removal of duplicates, two authors (A.H. and I.M.) independently screened the records for eligibility based on the titles and abstracts. Papers included at this stage were further appraised by reading the full text. Disagreement between reviewers was resolved by consulting a third reviewer (L.M.C.).

### Data collection

Using a preconstructed standardized data extraction form, two authors (A.H. and I.M.) independently collected data from the included records. From primary studies the following information was extracted (Table 1): first author’s name, year of publication, place of study, study type, population size, age, gender, method of diagnosis, type of PCR kit, outcome parameters: number of total, positive and negative saliva tests and number of total, positive and negative NPS test. From registered study protocols the following information was extracted and demonstrated in Table S2: Clinical Trial ID, Recruiting Status, Study type, Number of Centers and Study Design, Location, Population, Intervention, Comparison, Primary Outcomes, Secondary outcomes. In case of disagreement during extractions a third author (L.M.C.) was also involved.

**Table 1.** Summary of study characteristics of included records.

| First author and year | Country | Study type | Population |  | Diagnoses of COVID-19 | PCR kit | Reference standard | Index test | Outcome parameters |
| --- | --- | --- | --- | --- | --- | --- | --- | --- | --- |
|  |  |  | n (m/f) | Age |  |  |  |  |  |
| **Azzi et al. (2020)** | Italy | Consecutive case series | 25 (17/8) | 61 (mean) (39-85) | Viral RNA detection with PCR from NPS | Luna Universal qPCR Master Mix | NPS | Saliva | Number of positive and negative index tests |
| **Bae et al. (2020)** | South Korea | Consecutive case series | 4 (2/2) | 61.5 (35-82) | Viral RNA detection with PCR from NPS<br>And clinical signs of pneumonia | N/A | NPS | Saliva | Number of positive and negative index tests |
| **Fang et al. (2020)** | China | Consecutive case series | 32 (16/16) | 41 (34-54) | Viral RNA detection with PCR from NPS | N/A | NPS | Saliva | Number of positive and negative index tests |
| **To et al. (2020)** | Hong Kong, China | Consecutive case series | 23 (13/10) | 62 (37-75) | Viral RNA detection with PCR from NPS | QuantiNova Probe RT-PCR Kit | NPS | Saliva | Number of positive and negative index tests |
| **Williams et al. (2020)** | Australia | Consecutive case series | 39 (not published) | Not published | Viral RNA detection with PCR from NPS | Coronavirus Typing (835 well) assay | NPS | Saliva | Number of positive and negative index tests |
| **Not included in quantitative synthesis:** |  |  |  |  |  |  |  |  |  |
| **Deng and Hu (2020)** | China | Case report | 1 (0/1) | 39 | Viral RNA detection with PCR from NPS<br>And clinical signs of pneumonia | N/A | NPS | Saliva | Number of positive and negative reference tests and index tests |
| **Han et al. (2020)** | South Korea | Case report | 1 (0/1) | Neonate (27 day-old) | Viral RNA detection with PCR from NPS | PowerChek TM 2019-nCoV Real-time PCR Kit | NPS | Saliva | Number of positive and negative reference tests and index tests |
| **Wyllie et al. (2020)** | USA | Consecutive case series | 29 (16/13) | 59 (mean) (23-91) | Viral RNA detection with PCR from NPS | The US CDC real-time RT-PCR primer/probe sets | NPS | Saliva | Number of positive and negative reference tests and index tests |

NPS - Nasopharyngeal swab
N/A – Not available

### Risk of bias and applicability assessment

We evaluated the potential for bias, quality of reporting and applicability of the studies using the QUADAS-2 tool (Quality Assessment of Diagnostic Accuracy Studies 2) (Whiting, Rutjes, Westwood, Mallett, Deeks, Reitsma, Leeflang, Sterne, & Bossuyt, 2011), which is a tool widely used to assess studies of diagnostic accuracy. Our appraisal consisted of evaluating the risk of bias and applicability in four domains: 1) patient selection, 2) conduct and interpretation of index test and 3) reference standard, 4) flow and timing. We applied the following review question to judge their applicability to our investigation: *Are saliva specimens reliable for detecting SARS-CoV-2 in COVID-19 patients confirmed by nasopharyngeal swab testing?*

We used the preconstructed form available on the QUADAS-2 web page of the University of Bristol (Bristol).

### Summary measures and synthesis of results

In the synthesis of quantitative data we included patient-based data from consecutive case series. Case reports from single participants were excluded.

The sensitivity of the saliva test in the patient-based pooled data was calculated using the methods recommended by the working group of the Cochrane Collaboration. Because some of the sensitivity values are close to or equal to 1, the score confidence interval estimation (Wilson, 1927) was applied with the Freeman-Tukey double arcsine transformation (Freeman & Tukey, 1950). Due to the great variance in population size and methodologies, the random effect model by DerSimonian and Laird (DerSimonian & Laird, 1986) was used with 95% CI for random-effects meta-analysis.

Heterogeneity was assessed using the I^2^ measure and the ***χ***^2^ test, where p < 0.1 is taken to indicate significant heterogeneity. I^2^ values of 25%, 50%, and 75% were identified as low, moderate, and high estimates respectively. (Higgins, Altman, Gøtzsche, Jüni, Moher, Oxman, Savović, Schulz, Weeks, & Sterne, 2011). Statistical analyses were carried out using the STATA software version 15.0.

## Results

### Study selection

We included 20 articles for full-text evaluation of completed studies. Out of these, 8 were included in the qualitative synthesis, from which 5 were also included in the quantitative synthesis. Figure 1 illustrates the study selection process.

**Figure 1.**
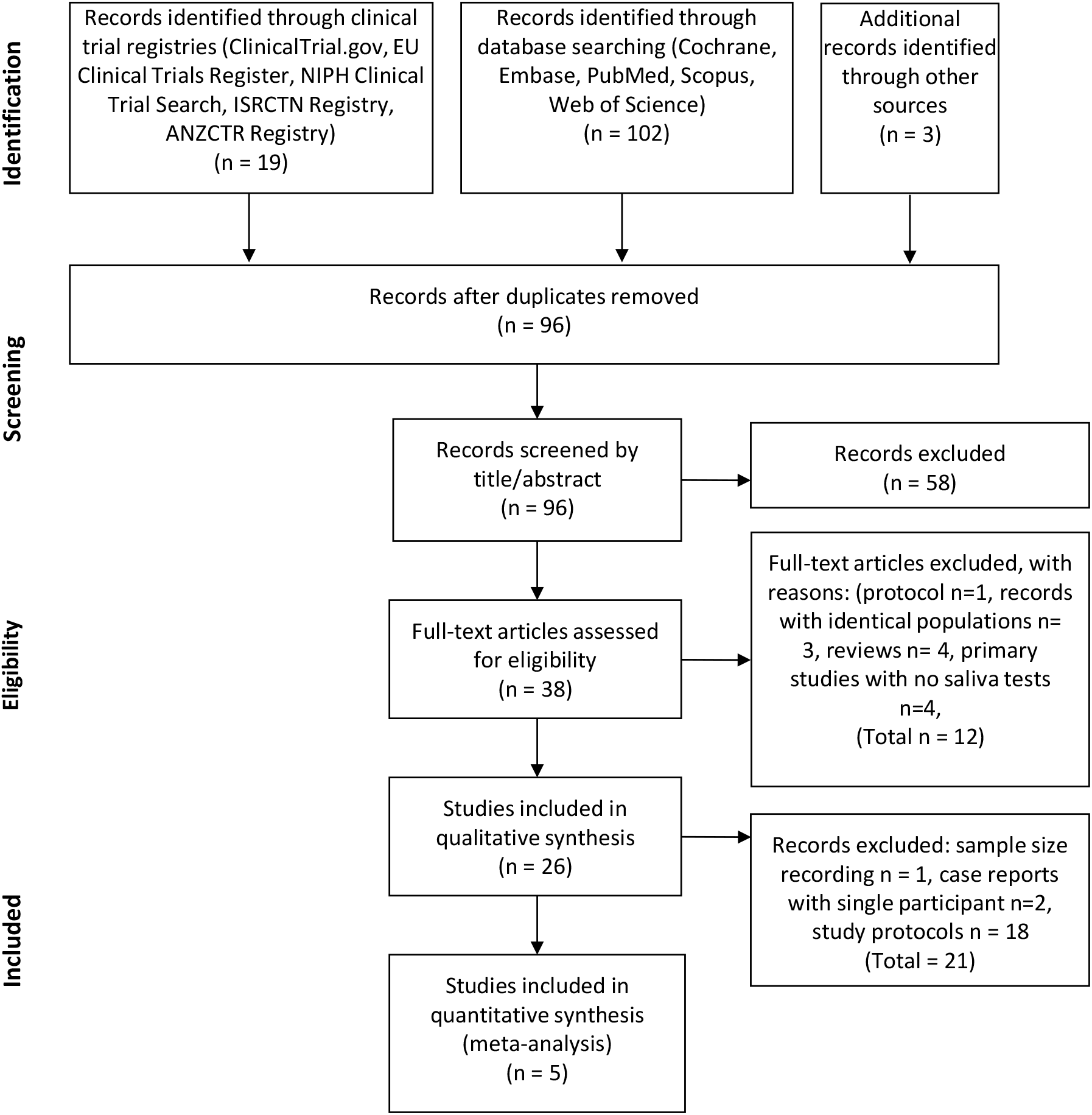
PRISMA flow diagram of the study selection process. Flow chart illustrating the selection process for identifying eligible records.

Our search in the clinical trial register yielded 19 protocols, out of which 1 was excluded due to different topic.

## Study characteristics

### Characteristics of the studies included

All five records included in the quantitative synthesis were consecutive case series, involving 123 patients from 5 distinct global locations (Table 1) (Azzi, Carcano, Gianfagna, Grossi, Gasperina, Genoni, Fasano, Sessa, Tettamanti, Carinci, Maurino, Agostino, Tagliabue, & Baj, 2020; Bae, Kim, Kim, Cha, Lim, Jung, Kim, Oh, Lee, Choi, Sung, Hong, Chung, & Kim, 2020; Fang, Zhang, Hang, Ai, Li, & Zhang, 2020; To, Tsang, Leung, Tam, Wu, Lung, Yip, Cai, Chan, Chik, Lau, Choi, Chen, Chan, Chan, Ip, Ng, Poon, Luo, Cheng, Chan, Hung, Chen, Chen, & Yuen, 2020; Williams, Bond, Zhang, Putland, & Williamson, 2020). All publications included patients with confirmed diagnoses of COVID-19. No other restrictions on inclusion were stated in any of the studies.

In the qualitative synthesis we also included another consecutive case series (Table 1). But, in their work Wyllie et al. presented 38 matching NPS and saliva samples from 29 patients without identifying double or multiple samplings of individual patients. Therefore, their sample-wise results cannot be combined for quantitative analysis with the others which reported patient-wise data (Wyllie, Fournier, Casanovas-Massana, Campbell, Tokuyama, Vijayakumar, Geng, Muenker, Moore, Vogels, Petrone, Ott, Lu, Lu-Culligan, Klein, Venkataraman, Earnest, Simonov, Datta, Handoko, Naushad, Sewanan, Valdez, White, Lapidus, Kalinich, Jiang, Kim, Kudo, Linehan, Mao, Moriyama, Oh, Park, Silva, Song, Takahashi, Taura, Weizman, Wong, Yang, Bermejo, Odio, Omer, Dela Cruz, Farhadian, Martinello, Iwasaki, Grubaugh, & Ko, 2020).

### Results of individual studies and synthesis of results

### Diagnostic potential of saliva specimens

In the individual studies included in the quantitative synthesis, the sensitivity of the saliva test among COVID-19 infected patients ranged from 78% (Fang et al., 2020) to 100% (Azzi et al., 2020).

Pooled event rates (positive and negative test results) from saliva specimens show that the sensitivity of the saliva test was 91% (CI 80% - 99%) among COVID-19 patients diagnosed in the recruitment period (Figure 2/A). Pooled event rates from NPS specimens taken during the studies after recruitment, in parallel to saliva specimen collections indicate that the sensitivity of the NPS test in these studies was 98% (CI 89% - 100%) (Figure 2/B). Since the two confidence intervals are overlapping, it appears that the positive test proportions of the saliva and NPS tests are not very different. However, it should be emphasized that this must be confirmed in the future when data will be available for diagnostic accuracy tests utilizing more clinical studies and 2×2 contingency tables.

**Figure 2.**
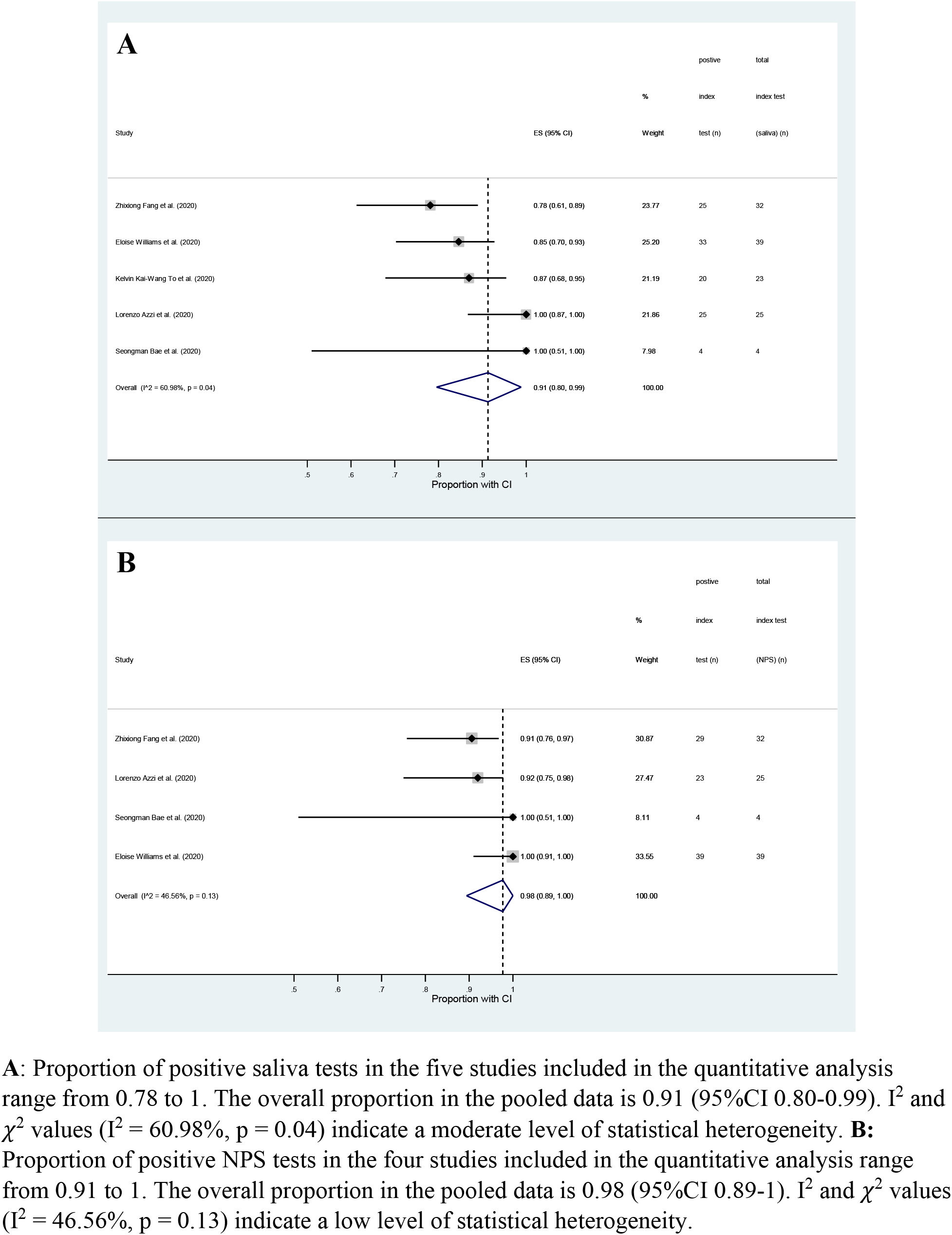
Meta-analysis of pooled event rates. **A**: Proportion of positive saliva tests in the five studies included in the quantitative analysis range from 0.78 to 1. The overall proportion in the pooled data is 0.91 (95%CI 0.80-0.99). I^2^ and *χ*^2^ values (I^2^ = 60.98%, p = 0.04) indicate a moderate level of statistical heterogeneity. **B:** Proportion of positive NPS tests in the four studies included in the quantitative analysis range from 0.91 to 1. The overall proportion in the pooled data is 0.98 (95%CI 0.89-1). I^2^ and *χ*^2^ values (I^2^ = 46.56%, p = 0.13) indicate a low level of statistical heterogeneity.

We evaluated our pooled results for inconsistency using the I^2^ test (Cumpston, Li, Page, Chandler, Welch, Higgins, & Thomas, 2019). In the case of salivary tests we found a moderate level of heterogeneity (I^2^ = 60.98%, p = 0.04) indicating the contribution of confounding factors in our analysis. On the other hand, we found a low level of heterogeneity among the NPS test results (I^2^ = 46.56 %, p = 0.13).

Interestingly some of the data suggest that NPS tests may occasionally be negative when the saliva test gives a positive result. In the study of Wyllie et al. the viral RNA in 8 patients was detected only in their saliva (Wyllie et al., 2020). Azzi et al. reported that two patients showed positive saliva tests while their NPS tests were negative (Azzi et al., 2020). And a case report showed that in seven samples from one individual there was no NPS positivity while the saliva specimen was positive (Deng, Hu, Yang, Zheng, Peng, Ren, Zeng, & Tian, 2020).

In a more detailed study, Bae et al. examined the difference in viral loads between the two sampling methods; the values ranged from 0.06 to 3.39 log_10_ units higher in the NPS specimens than in the saliva specimens (Bae et al., 2020). One case series (Williams et al., 2020) and another case report on a 27-day-old neonate (Han, Seong, Heo, Park, Kim, Shin, Cho, Park, & Choi, 2020) also found that there were higher viral loads in the NPS specimens. On the other hand, in a sample-based study Wyllie et al. (Wyllie et al., 2020), using 38 matched samples, detected SARS-CoV-2 in saliva but not in 8 NPS samples (21%), while detected SARS-CoV-2 in NPS and not saliva only in 3 matched samples (8%). Furthermore, they found significantly higher SARS-CoV-2 titers from saliva than NPS. Unfortunately, they did not present patient-based matched data, therefore, these observations could not be involved in our above described quantitative statistical analysis.

Only two study assessed the specificity of saliva tests besides sensitivity (Williams et al., 2020) (Wyllie et al., 2020). In one work a subset of saliva specimens from 50 patients with PCR-negative swabs was tested. SARS-CoV-2 was detected in 1/50 (2%; 95% CI 0.1%-11.5%) of these saliva samples Williams, 2020 #467}. The other tested 98 asymptomatic healthcare workers with parallel NPS and saliva tests. NPS tests turned out to be negative for all participants, while saliva tests were positive for two (Wyllie et al., 2020).

### Risk of bias within studies

We assessed risk of bias in the six included case series (Azzi et al., 2020; Bae et al., 2020; Fang et al., 2020; To et al., 2020; Williams et al., 2020; Wyllie et al., 2020) according to the QUADAS-2 tool. Five studies (Azzi et al., 2020; Bae et al., 2020; Fang et al., 2020; To et al., 2020; Wyllie et al., 2020) had low risk of bias in selection bias. On the other hand 4 studies (Azzi et al., 2020; Bae et al., 2020; Fang et al., 2020; To et al., 2020) had high risk of bias in the index test due to the fact that the saliva tests results were interpreted with the knowledge of the results of the reference standard. Flow and timing were high or unclear in all studies, since there were no exact information regarding the time passed between specimens collection for the two tests. Applicability had low concerns in index test in four (Azzi et al., 2020; To et al., 2020; Williams et al., 2020; Wyllie et al., 2020) and unclear in two studies (Bae et al., 2020; Fang et al., 2020). The summary of the risk-of bias analysis and applicability concerns is available in Table S2 and in Table S3.

### Ongoing registered clinical trials on saliva diagnostics for COVID-19

We systematically searched for clinical trial protocols that are planning to evaluate saliva specimens for COVID-19 diagnosis in 5 clinical trial registers (EU Register, ISRCTN, ANZCTR, JPRN, ClinicalTrials.gov). By using the same keywords as we applied for already completed studies, we found 18 registered clinical trials on planned or ongoing clinical studies. All of them appeared in the registry ClinicalTrials.gov (Table S2). Among these, 13 studies are noninterventional. These investigations primarily focus on the diagnostic values of various specimens collected from patients, including NPS, saliva, blood and others to identify the diagnostic and prognostic values of such samples in detecting and following the progression of COVID-19 disease. The additional 5 interventional studies are examining the effectiveness of several potentially beneficial compounds, such as azithromycin, lopinavir/ritonavir, beta-cyclodextrin, citrox 3 and peginterferon lambda on the outcomes of viral infection. In these studies, besides NPS specimen collections, saliva tests are also planned. In the trial protocols very little information is available about the optimization and validation of saliva collection, transportation and storage of saliva samples, nor about the viral RNA assay methods to be used for saliva samples, and the choice of appropriate internal controls in view of the scarcity of human DNA in saliva samples.

## Discussion

In April 2020 the Food and Drug Administration (FDA) granted emergency use authorization (EUA) to Rutgers’ RUCDR Infinite Biologics and its collaborators for a new specimen collection approach that utilizes saliva as the primary test biomaterial for the SARS-CoV-2 coronavirus, the first such approval granted by the federal agency (https://www.fda.gov/media/136877/download). This new saliva-based diagnostic collection method, which RUCDR has developed in partnership with Spectrum Solutions and Accurate Diagnostic Labs (ADL), claims to allow an easier and therefore broader screening of the population compared with the current method using nose and throat swabs. Another accelerated EUA for the “Curative-Korva SARS-Cov-2 Assay” was also approved to permit the testing of oral fluids, i.e. saliva (https://www.fda.gov/media/137088/download). This assay was specifically designed for use with oral fluid specimens. Nasopharyngeal swabs, oropharyngeal swabs and nasal swabs can also be used with the Curative-Korva SARS-CoV-2 Assay, but their performance with this assay has not yet been assessed (https://www.fda.gov/media/137088/download). These two saliva-based, FDA-approved assays are now in intensive use to test for COVID-19 infection, in spite of the fact that no independent, scientific analysis has not yet established their effectiveness. Our present work is the first integrative meta-analysis study to review the existing multi-study evidence for the saliva-based approach.

The use of saliva as a diagnostic tool for various systemic conditions is nothing new. Considerable research effort has been made in the past to seek biomarkers in saliva, since its collection is noninvasive and easy. As a result, emerging evidence indicates that whole saliva can be used to identify various oral and systemic conditions (for reviews see (Dawes & Wong, 2019; Keremi et al., 2017) (Kaczor-Urbanowicz, Martin Carreras-Presas, Aro, Tu, Garcia-Godoy, & Wong, 2017)). Importantly, the concept of using saliva to detect viral infections is now well established (Niedrig et al., 2018) (Corstjens, Abrams, & Malamud, 2012)).

Among RNA viruses, salivary diagnostic tests for Zika are well elaborated ((Khurshid, Zafar, Khan, Mali, & Latif, 2019) (Gorchakov, Berry, Patel, El Sahly, Ronca, & Murray, 2019).) and a number of salivary-based detection methods have been reported for Ebola virus detection (Niedrig et al., 2018). The presence of considerable quantities of viral RNA in the saliva of 17 SARS-infected patients has also been shown unequivocally (Wang, Chen, Liu, Chen, Chen, Yang, Chen, Yeh, Kao, Huang, Hsueh, Wang, Sheng, Fang, Hung, Hsieh, Su, Chiang, Yang, Lin, Hsieh, Hu, Chiang, Wang, Yang, & Chang, 2004). But most studies lack any direct comparison of the sensitivity and specificity of NPS- and saliva-based assays. The one important exception is a study which compared saliva and NPS specimens for the detection of respiratory viruses by multiplex RT-PCR (Kim et al., 2017). This study, which included results from 236 patients with 11 different viral respiratory infections, including coronaviruses, revealed no significant difference in the sensitivity and specificity of saliva- and NPS-based tests (Kim et al., 2017). Taken together, although saliva-based diagnostics are supported by a considerable amount of evidence, routine applications are still rare because of the lack of well standardized protocols.

The source of SARS-CoV-2 in saliva is unknown at present but it could come from multiple locations. One obvious source is debris from the nasopharyngeal epithelium which drains into the oral cavity (To et al., 2020). Secondly, SARS-CoV-2 may actually infect the salivary glands and the virus is then secreted into the saliva from the glands. No information is available on this. But it is of note that during the infection of rhesus macaques by the SARS coronavirus, epithelial cells lining salivary gland ducts are an early target of the virus (Liu, Wei, Alvarez, Wang, Du, Zhu, Jiang, Zhou, Lam, Zhang, Lackner, Qin, & Chen, 2011). One consequence of this is the production of SARS-specific secretory immunoglobulin A into the saliva (Lu, Huang, Huang, Li, Zheng, Chen, Chen, Hu, & Wang, 2010). Thirdly, SARS from blood plasma may access the mouth via the crevicular fluid, an exudate derived from periodontal tissues (Silva-Boghossian, Colombo, Tanaka, Rayo, Xiao, & Siqueira, 2013). Fourthly, infected oral mucosal endothelial cells, which show overexpression of ACE2 during SARS-CoV-2 infection may also contribute to viral load in saliva (Xu, Zhong, Deng, Peng, Dan, Zeng, Li, & Chen, 2020). Finally, salivary cells may endocytose viruses and virus-containing exosomes from the circulation at their basolateral surface and release them into the salivary lumen by exocytosis. Such mechanisms have been revealed for other macromolecular constituents of the blood, such as DNA and RNA in exosomes (Dawes & Wong, 2019). Any or all of these five possible sources may contribute to the appearance of SARS-CoV-2 in the saliva of COVID-19 patients.

In the present meta-analysis we found that the test sensitivities were 91% (CI 80% - 99%) and 98% (CI 89% - 100%) for saliva and for NPS samples, respectively, based the pooled event rates among COVID-19 patients. Clearly, the two confidence intervals overlap, suggesting that the outcomes of the saliva tests and NPS tests are not very different, although a tendency for NPS to be more sensitive is numerically visible. On the other hand, one study, which could not be included in the main quantitative analysis because it used a different sampling protocol, reported the opposite tendency. On a significant number of occasions (21%) they detected SARS-CoV-2 in saliva but not in matched NSP samples, whereas SARS-CoV-2 was detected in NSP and not saliva on just three occasions (8%) (Wyllie et al., 2020). Although NPS-based SARS-CoV-2 virus detection is currently regarded as the gold standard (Lippi et al., 2020; Sullivan, Sailey, Guest, Guarner, Kelley, Siegler, Valentine-Graves, Gravens, Del Rio, & Sanchez, 2020; Zou, Ruan, Huang, Liang, Huang, Hong, Yu, Kang, Song, Xia, Guo, Song, He, Yen, Peiris, & Wu, 2020), carefully performed future studies need to be carried out to determine both the sensitivity and specificity of the NPS and saliva tests in parallel measurements to firmly establish the relative diagnostic accuracies of these applying 2×2 contingency tables for statistical analysis.

At present only two study have assessed the specificity of the saliva tests. In one of n those tests only one saliva sample was found to be positive among 50 apparently healthy individuals who were PCR-negative for the NPS test (Williams et al., 2020). In the other work two individuals were detected positive using saliva tests among 98 participants who were negative for NPS test (Wyllie et al., 2020). This results may reflect a real difference in the specificities of the NPS and saliva tests.

For optimal saliva-based testing at least three conditions have to be improved by standardization then validation (Bhattarai, Kim, & Chae, 2018). 1) A specific saliva collection method should be selected and optimized after sytematically comparing the various methods currently used for collecting whole saliva. 2) The optimal solution for collecting, transporting and storing saliva samples should be found. 3) The RNA assay method, either RT-PCR or loop mediated isothermal amplification (LAMP) or another protocol, should also be optimized for saliva, using an appropriate internal control; this cannot be human DNA which is overwhelming in NPS but not in saliva samples (Bae et al., 2020; Fang et al., 2020; To et al., 2020; Williams et al., 2020; Wyllie et al., 2020).

The studies included in our analysis used different sampling methods to collect saliva. This may have had a significant effect on the sensitivity of the saliva test. Azzi et al. used a simple drooling technique to collect saliva and they resuspended the collected specimens in 2 ml of PBS. In contrast, To et al. 2020 collected saliva specimens containing fluid from the posterior oropharynx obtained by coughing up and clearing the throat (To et al., 2020). Another study (Williams et al., 2020) asked patients to pool saliva in their mouth prior to collection, and to spit 1-2 ml into a collection pot. The act of pooling saliva in the mouth may have stimulated additional saliva secretion, which could have diluted the specimen. In this case no transport medium was added to the specimens but, after transportation to the laboratories, liquid Amies medium was added. Wyllie et al. used a self-collection technique: patients were asked to spit repeatedly into a sterile urine cup until one third was full. This too could have diluted the sample with additional virus-free saliva. The remaining two studies did not describe the collection method at all (Bae et al., 2020; Fang et al., 2020). Additionally, two of the studies specified that specimens were collected in early morning to avoid eating, drinking and tooth brushing (To et al., 2020; Wyllie et al., 2020).The rest of the studies did not specify the time of collection or mention any confounding factors that may have affected the sample.

Other factors, such as the type of transport medium, temperature during transportation, time passed between specimen collection and RNA extraction may also affect the outcome of the tests (Bhattarai et al., 2018). Unfortunately, there is insufficient information in these few studies to draw any conclusions about the possible effects of these confounding factors on the accuracy of saliva testing for COVID-19 diagnosis.

It is likely that the simple drooling technique, with no specific target volume, will provide the greatest sensitivity if the viral RNA in whole saliva derives from sources other than the secretions of the salivary glands. Drooling is a well-established saliva collection method that is generally recommended for analytical purposes (Golatowski, Salazar, Dhople, Hammer, Kocher, Jehmlich, & Volker, 2013). Due to its simplicity, it does not require trained personnel and can even be self-administered. Additionally, the drooling method is much safer. Saliva is drooled directly into a container from the mouth, with no need for infected swabs to be carried in the air from patient’s nostril to the container therefore reducing the risk to healthcare staff. Moreover, this saliva collecting technique is suitable to avoid the mixing of fluids from different anatomical regions as well (e.g. oropharynx), since it only collects fluid from the oral cavity (Azzi et al., 2020).

The need for reliable, non-invasive and easy-to-perform tests for COVID-19 has triggered special attention in the last few months. Between 1 January and 25 April 2020 the commencement of 18 clinical trials have been reported in the ClinicalTrials.gov registry for saliva specimens (Table S2). Among these, 13 studies are non-interventional and these focus on the diagnostic values of various specimens including saliva. Five interventional studies also plan to use saliva as a diagnostic tool together with NPS specimens, but their primary focus is on evaluating potential treatments for SARS-CoV-2 infections. Unfortunately, these registered clinical trials vary considerably in the amount of information presented about the testing methodology. Neither the non-interventional, nor the interventional protocols have clear descriptions of the collection, transportation and storage of saliva samples, and the optimization of viral RNA assays suitable for saliva specimens. Only a few of them emphasize the necessity for determining the sensitivity and specificity of the saliva-based test. But hopefully, during the course of execution, such studies will yield high quality, reliable data on that matter.

## Limitations

A limitation of the present work is the relatively small number of studies and small sample sizes available regarding this topic. Despite the large number of records found by the systematic search, only 6 could be included. Although intensive research is on progress regarding COVID-19, there are only a handful articles fulfilling our eligibility criteria. The limited number of reported data makes it difficult to perform comprehensive analyses and to thoroughly investigate the causes behind certain trends. Another issue that hinders in-depth analysis is the inhomogeneous methodologies, insufficient and deficient reporting of methods and outcome parameters. A significant limitation is the lack of data for 2×2 contingency tables since almost no specificity data are available as yet. Thus, accurate statistical methodologies (such as use of a bivariate model) specially developed for meta-analysis of diagnostic test accuracy could not be used in this work All studies except two (Williams et al., 2020; Wyllie et al., 2020), investigated the reliability of saliva test only among COVID-19 infected participants, no healthy individuals were recruited. Additionally, there are several other confounding factors which might affect the detectability of viral RNA from saliva, such as time of sample collection, method for saliva collection, virus transport medium, storage and transport temperatures, time passed between specimen collection and RNA isolation, extraction kits and PCR kits used for isolation, amplification and detection. Due to limited data the potential effect these parameters could not be investigated in our analysis.

## Conclusion

In the present meta-analysis we provide evidence that saliva tests are a promising alternative to nasopharyngeal swabs for COVID-19 diagnosis. Optimized and validated saliva assays may provide the possibility of reliable self-collection of samples for COVID-19 testing in the future. However, there are many open questions to be answered for the specificity and sensitivity of saliva-based tests. Therefore, much more research is needed in order to routinely introduce determination of SARS-CoV-2 using saliva specimens in clinical practice.

## Data Availability

The data that support the findings of this study are available from the corresponding author (G.V.), upon reasonable request.

## Legends for Supplementary files

**Table S1.**
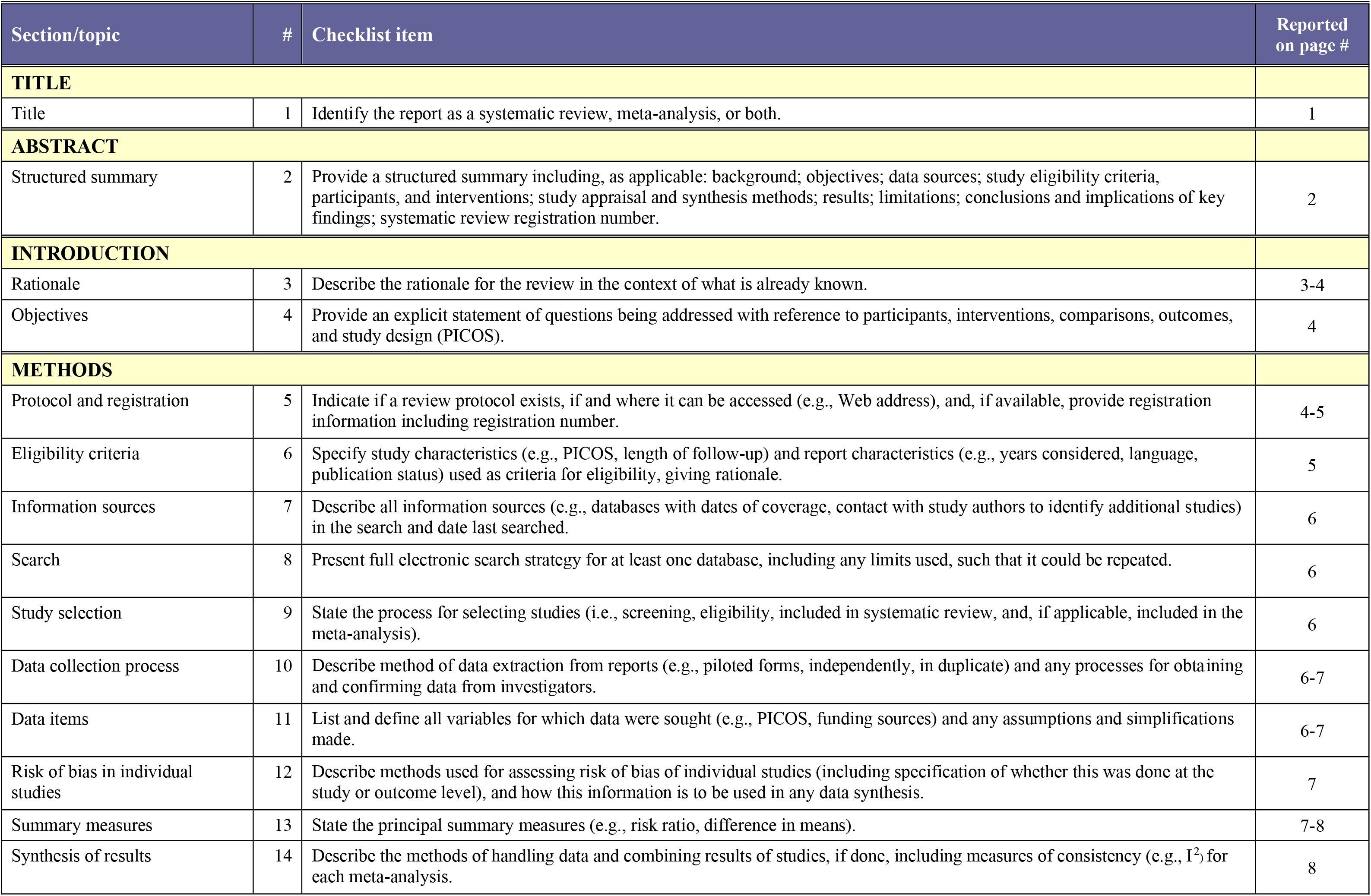
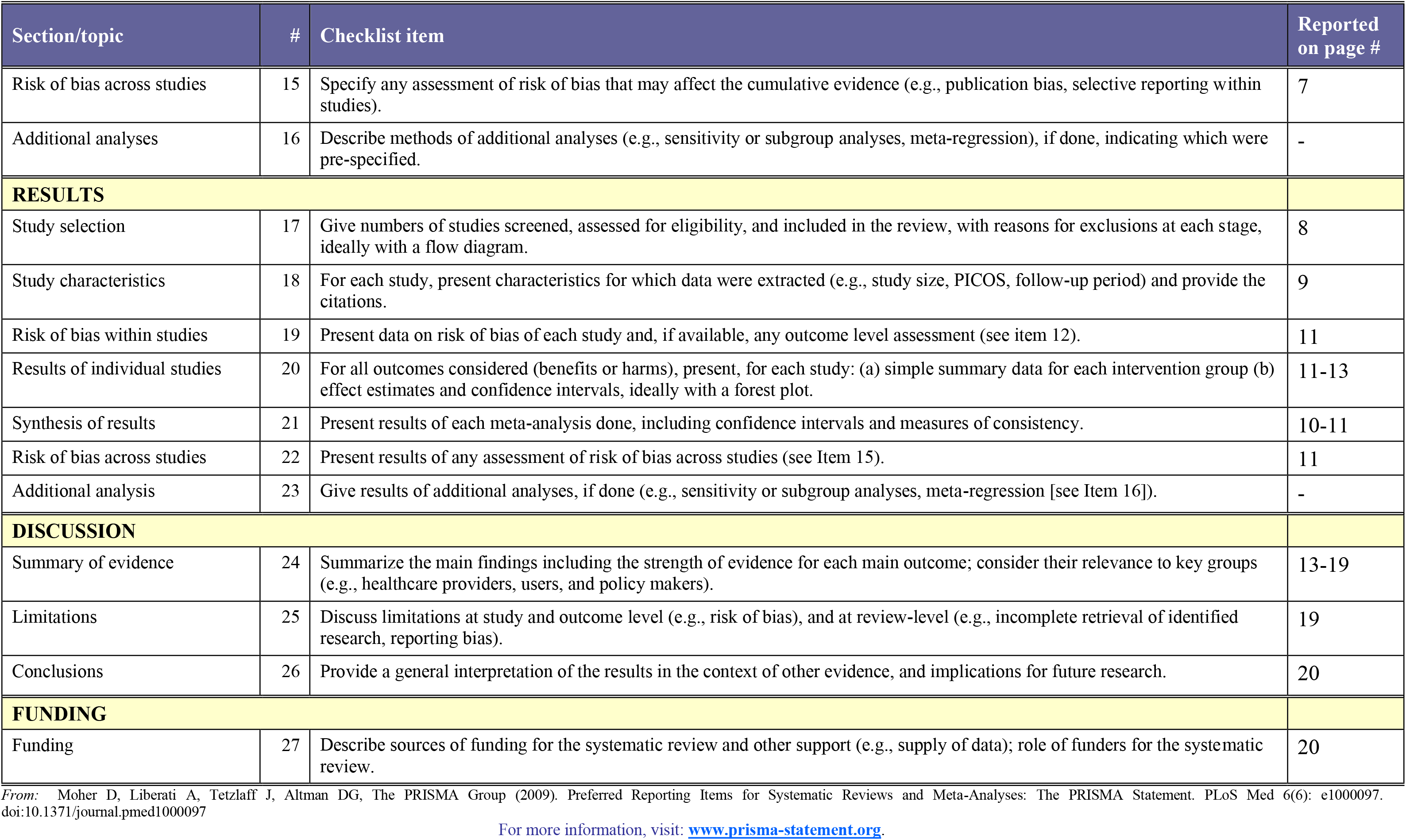
PRISMA checklist

**Table S2.** Characteristics of clinical trials including saliva as a diagnostic tool for COVID-19, registered on ClinicalTrials.gov

| **ID** | NCT04332107 | NCT04321174 | NCT04352959 | NCT04354259 | NCT04276688 |
| --- | --- | --- | --- | --- | --- |
| **Recruiting Status** | Recruiting | Recruiting | Recruiting | Recruiting | Completed |
| **Study type** | interventional | interventional | interventional | interventional | interventional |
| **Number of Centers and Study Design** | Single center, interventional, randomized (RCT), parallel assignment, quadruple masking, phase 3 | Multi-locations, interventional, randomized (RCT), parallel assignment, single masking (outcomes assessor), phase 3 | Multi-locations, interventional, randomized (RCT), parallel assignment, triple masking | Single center, interventional, randomized (RCT), parallel assignment, none masking, phase 2 | Single center, interventional, randomized (RCT), parallel assignment, none masked, Phase 2 |
| **Location** | the USA | Canada | France | Canada | China |
| **Population** | Subjects with positive SARS-CoV-2 test results received within the previous three days, but not hospitalized (n=2271) | 1) High risk close contact with a confirmed COVID-19 case during their symptomatic period, 2) Successfully contacted by the study team within 24 hours of study team notification of the relevant index COVID-19 case (n=1220) | Patients with clinical diagnosis of Covid-19 infection (n=178) | 1) For ambulatory cohort: patients confirmed COVID-19 infection by PCR within 5 days of symptom onset discharged to home isolation, 2) For hospitalized cohort: SARS-CoV-2 RNA-positive on nasopharyngeal swab / respiratory specimen within 5 days of symptom onset admitted to hospital for management of COVID-19 (n=140) | Subjects include patients hospitalized for confirmed 2019-n-CoV infection, temperature $\geq 38^{\circ}\text{C}$ with another symptoms upon admission (n=127) |
| **Intervention** | Single oral 1g dose of Azythromycin | Lopinavir/Ritonavir 400/100 mg twice daily for 14 days | Mouthrinse with bêta-cyclodextrin and citrox 3 daily mouthrinses for 7 days | Single dose of peginterferon lambda 180$\mu\text{g}$ sc at baseline for ambulatory cohort and peginterferon lambda 180$\mu\text{g}$ sc at baseline and a second dose on day 7 for hospitalized cohort | Lopinavir/Ritonavir 400/100 mg twice daily for 14 days, Ribavirin 400 mg twice daily for 14 days and IFN-beta-1B 0.25 mg sc injection alternate day for 3 day / Nasopharyngeal swab, saliva, urine, stool and blood sampling |
| **Comparison** | placebo | no intervention | Placebo: mouth rinse without antiviral | No specific therapy for ambulatory cohort and the best supportive care for hospitalized cohort | Lopinavir/Ritonavir 400/100 mg twice daily for 14 days |
| **Primary Outcomes** | All-cause hospitalization or emergency room stay of >24 hours | The primary outcome is microbiologically confirmed COVID-19 infection, ie. detection of viral RNA in a respiratory specimen (mid-turbinate | Change from baseline amount of SARS-CoV-2 in salivary samples at 7 days | 1) The proportion of participants with negative SARS-CoV-2 RNA on nasopharyngeal swab, Nasopharyngeal swab, saliva and blood sampling. 2) Rate of combined treatment-emergent | Time to negative nasopharyngeal swab |
|  |  | swab, nasopharyngeal swab, sputum specimen, saliva specimen, oral swab, endotracheal aspirate, bronchoalveolar lavage specimen) by day 14 of the study. |  | and treatment-related severe adverse events |  |
| **Secondary outcome** | viral load by self-collected nasal swab, Viral load by self-collected saliva swab | - | Change from Baseline amount of SARS-CoV-2 virus in nasal samples at 7 days | - | Time to negative saliva 2019-n-CoV RT-PCR |

**Table S2.** Continued
| **ID** | NCT04360811 | NCT04354610 | NCT04361604 | NCT04325919 | NCT04351646 |
| --- | --- | --- | --- | --- | --- |
| **Recruiting Status** | Recruiting | Recruiting | Not yet recruiting | Recruiting | Recruiting |
| **Study type** | non-interventional | non-interventional | non-interventional | non-interventional | non-interventional |
| **Number of Centers and Study Design** | Single center, observational, non-randomized (NRCT), parallel assignment, none masked | Multi-locations, observational, single group assignment, none masked | Single center, observational, cohort, prospective | Observational | Single center, observational, case-control, prospective |
| **Location** | France | France | France | China | UK |
| **Population** | 1) Unexposed group: COVID 19 negative pregnant woman, 2) Exposed group: COVID 19 positive (symptomatic and asymptomatic) pregnant woman (n=3600) | Patients hospitalized for critical form of Covid-19 infection within 3 days (n=57) | 1) Patients co infected HIV and SRAS-CoV2 (n=250), 2) Patients infected HIV without COVID-19 (n=20) | Patients with laboratory-confirmed COVID-19, (n=170) Patients hospitalized for pneumonia tested negative for COVID-19 are controls | 1) SARS-CoV-2 negative inpatients, 2) SARS-CoV-2 positive inpatients, 3) SARS-CoV-2 suspected or confirmed SARS-CoV-2 positive cases amongst health care professionals and lab staff (n=500) |
| **Intervention** | N/A | N/A | N/A | N/A | N/A |
| **Comparison** | N/A | N/A | N/A | N/A | N/A |
| **Primary Outcomes** | Exposure to SARS-CoV-2 will be measured the day of delivery by RT-PCR on maternal saliva and by serology on maternal blood | 1) Worsening of renal function by at least KDIGO grade 1 during hospitalization for Covid-19 infection, 2) Troponin greater than 99th percentile during hospitalization for Covid-19 infection | Describe the course of COVID-19 disease in patients infected with HIV, biological sampling (blood, saliva, rectal swab (stool swab), urine, nasopharyngeal swab, conjunctival swab, semen | Patients' treatment and management during hospitalization. Serial viral load changes during hospitalization. Collection of blood, stool, rectal swab, urine, saliva, nasopharyngeal aspirate/flocked swab, sputum/tracheal aspirate | Antibody titres to SARS-CoV-2 at specified days post baseline samples (Nasopharyngeal swab, blood and saliva sampling) |
| **Secondary outcome** | Description of the number of positive COVID-19 RT-PCRs in the conception products: amniotic fluid, frozen placenta fragment, frozen fetal tissue, cord blood or frozen cord fragment | Blood samples, **saliva** collection, and urine collection to carry out biomarker assays and for the constitution of a biological collection. | - | - | - |

**Table S2.** Continued
| **ID** | NCT04337424 | NCT04357977 | NCT04356586 | NCT04355533 | NCT04362150 |
| --- | --- | --- | --- | --- | --- |
| **Recruiting Status** | Recruiting | Recruiting | Enrolling by invitation | Recruiting | Recruiting |
| **Study type** | non-interventional | non-interventional | non-interventional | non-interventional | non-interventional |
| **Number of Centers and Study Design** | Single center, observational, case-control, prospective | Multi-locations, observational, cross-sectional | Single center, observational, cohort, prospective | Single center, observational, non-randomized (NRCT), single group assignment, none masked | Single center, observational, cohort, prospective |
| **Location** | France | USA | Belgium | France | USA |
| **Population** | 1) Patients diagnosed positive,<br>2) Healthcare staff presumed negative for SARS-CoV-2 (n=180) | Patients and study staff at the testing site who have been flagged for COVID-19 testing or who are being treated for COVID-19 (n=300) | Healthcare workers with mild symptoms for Covid-19 (n=300) | Children hospitalized since at most 4 days and their parents (n=1920) | Individuals with positive test for COVID-19 who have recovered from acute infection (wide spectrum of age, race, gender and disease severity) (n=800) |
| **Intervention** | N/A | N/A | N/A | N/A | N/A |
| **Comparison** | N/A | N/A | N/A | N/A | N/A |
| **Primary Outcomes** | Comparison of LAMP test with reference RT-PCR on viral detection (Saliva and nasopharyngeal swab sampling) | RBA-2 saliva monitoring device development. Nasopharyngeal swab and saliva sample. The comparison of the results obtained from the current testing methods will be used to calibrate machine learning algorithms of the RBA-2 | 1) Percentage of serological positive healthcare workers, 2) Percentage of healthcare workers with positive saliva swabs | Seroconversion against SARS-CoV2 in children, Nasopharyngeal, rectal swabs, saliva and blood sampling | Demographic data on participants and Proportion of participants previously hospitalized. Whole blood, peripheral blood mononuclear cells, plasma, serum and saliva. |
| **Secondary outcome** | - | - | - | - | - |

**Table S2.** Continued
| **ID** | NCT04357327 | NCT04336215 | NCT04348240 |
| --- | --- | --- | --- |
| **Recruiting Status** | Recruiting | Recruiting | Recruiting |
| **Study type** | non-interventional | non-interventional | non-interventional |
| **Number of Centers and Study Design** | Single center, non-randomized (RCT), parallel assignment, single masking | Multi-locations, observational, cohort, prospective | Single center, observational, cohort, prospective |
| **Location** | Italy | USA | USA |
| **Population** | 1) Patients with symptoms associated with COVID-19, 2) Asymptomatic patients with low risk phenotype (n=100) | 1) Healthcare workers (n=500), 2) Non-healthcare workers: faculty staff and students, who do not have patient contact (n=250), 3) Multigenerational household members, who test positive and negative for SARS-CoV-2 (n=540) | 1) Asymptomatic high-risk subjects with known history of close personal contact with a COVID-19 positive person not tested (SARS-CoV2 status unknown), 2) Asymptomatic or mildly symptomatic subjects who are COVID-19 positive, 3) COVID-19 positive individuals retesting negative (n=60) |
| **Intervention** | N/A | N/A | N/A |
| **Comparison** | N/A | N/A | N/A |
| **Primary Outcomes** | 1) Sensibility after 10 minutes for salivary test and after 6 hours for the nasopharyngeal swab, 2) Specificity after 10 minutes for salivary test and after 6 hours for the nasopharyngeal swab | 1) Prevalence, 2) Incidence, Nasopharyngeal swab, saliva and blood sampling | Determination of SARS-CoV-2 viral load and infectivity in saliva that may contribute to asymptomatic transmission. Collection of nasal and oral secretions and droplets produced by participants while they speak |
| **Secondary outcome** | - | - | - |

**Table S3.** Summary of risk-of-bias and applicability concerns in included studies.

| STUDY | RISK OF BIAS |  |  |  | APPLICABILITY CONCERNS |  |  |
| --- | --- | --- | --- | --- | --- | --- | --- |
|  | PATIENT SELECTION | INDEX TEST | REFERENCE STANDARD | FLOW AND TIMING | PATIENT SELECTION | INDEX TEST | REFERENCE STANDARD |
| Azzi et al. (2020) | ✓ | ✗ | ✓ | ✗ | ✓ | ✓ | ✓ |
| Bae et al. (2020) | ✓ | ✗ | ✓ | ? | ✓ | ? | ? |
| Fang et al. (2020) | ✓ | ✗ | ✓ | ? | ✓ | ? | ✓ |
| To et al. (2020) | ✓ | ✗ | ? | ? | ✓ | ✓ | ✓ |
| Williams et al. (2020) | ? | ? | ? | ✗ | ✓ | ✓ | ? |
| Not included in the quantitative analysis: |  |  |  |  |  |  |  |
| Wyllie et al. (2020) | ✓ | ? | ? | ? | ✓ | ✓ | ✓ |

✓ = Low Risk ✗ = High Risk ? = Unclear Risk

**Table S4.** Detailed summary of risk of bias and applicability across studies.

| **<u>Risk of bias</u>** | **Yes** | **No** | **Unclear** |
| --- | --- | --- | --- |
| **DOMAIN 1: PATIENT SELECTION** |  |  |  |
| Was a consecutive or random sample of patients enrolled? | 6 | 0 | 0 |
| Was a case-control design avoided? | N/A |  |  |
| Did the study avoid inappropriate exclusions? | 5 | 0 | 1 |
| **Could the selection of patients have introduced bias?** | **5** | **0** | **1** |
| **DOMAIN 2: INDEX TEST(S)** |  |  |  |
| Were the index test results interpreted without knowledge of the results of the reference standard? | 0 | 4 | 2 |
| If a threshold was used, was it pre-specified? | 0 | 0 | 6 |
| **Could the conduct or interpretation of the index test have introduced bias?** | **0** | **4** | **2** |
| **DOMAIN 3: REFERENCE STANDARD** |  |  |  |
| Is the reference standard likely to correctly classify the target condition? | 4 | 0 | 2 |
| Were the reference standard results interpreted without knowledge of the results of the index test? | 3 | 0 | 3 |
| **Could the reference standard, its conduct, or its interpretation have introduced bias?** | **3** | **0** | **3** |
| **DOMAIN 4: FLOW AND TIMING** |  |  |  |
| Did all patients receive a reference standard? | 6 | 0 | 0 |
| Did patients receive the same reference standard? | 6 | 0 | 0 |
| Were all patients included in the analysis? | 5 | 1 | 0 |
| **Could the patient flow have introduced bias?** | **0** | **2** | **4** |
| **<u>Applicability concerns</u>** | **Low** | **High** | **Unclear** |
| **DOMAIN 1: PATIENT SELECTION** |  |  |  |
| **Is there concern that the included patients do not match the review question?** | **6** | **0** | **0** |
| **DOMAIN 2: INDEX TEST(S)** |  |  |  |
| **Is there concern that the index test, its conduct, or interpretation differ from the review question?** | **4** | **0** | **2** |
| **DOMAIN 3: REFERENCE STANDARD** |  |  |  |
| **Is there concern that the target condition as defined by the reference standard does not match the review question?** | **4** | **0** | **2** |

